# Vision Transformer Autoencoders for Unsupervised Representation Learning: Revealing Novel Genetic Associations through Learned Sparse Attention Patterns

**DOI:** 10.1101/2025.03.24.25324549

**Authors:** Samia R. Islam, Wei He, Ziqian Xie, Degui Zhi

**Affiliations:** D. Bradley McWilliams School of Biomedical Informatics The University of Texas Health Science Center at Houston

**Keywords:** vision transformer, deep learning, brain MRI, medical imaging, GWAS

## Abstract

The discovery of genetic loci associated with brain architecture can provide deeper insights into neuroscience and potentially lead to improved personalized medicine outcomes. Previously, we designed the Unsupervised Deep learning-derived Imaging Phenotypes (UDIPs) approach to extract phenotypes from brain imaging using a convolutional (CNN) autoencoder, and conducted brain imaging GWAS on UK Biobank (UKBB). In this work, we design a vision transformer (ViT)-based autoencoder, leveraging its distinct inductive bias and its ability to capture unique patterns through its pairwise attention mechanism. The encoder generates contextual embeddings for input patches, from which we derive a 128-dimensional latent representation, interpreted as phenotypes, by applying average pooling. The GWAS on these 128 phenotypes discovered 10 loci previously unreported by CNN-based UDIP model, 3 of which had no previous associations with brain structure in the GWAS Catalog. Our interpretation results suggest that these novel associations stem from the ViT’s capability to learn sparse attention patterns, enabling the capturing of non-local patterns such as left-right hemisphere symmetry within brain MRI data. Our results highlight the advantages of transformer-based architectures in feature extraction and representation learning for genetic discovery.

## Introduction

Insights into understanding brain structural anatomy and related genetic signals have the potential to pave the way for breakthroughs in neuroscience and precision medicine. Most genome-wide association studies (GWAS) in the past have focused on direct phenotype measurements, such as brain region volumes, cortical surface areas, and cortical thickness, estimated using traditional software tools like FSL and FreeSurfer^1–5^, or statistical and mathematical representations of anatomical modeling and population genetics^6–11^. While these approaches have led to interesting discoveries, they rely on prior knowledge and constraints and may overlook more nuanced or complex patterns in the data.

Recent advances in machine learning techniques offer more flexible and data-driven methods that can learn important features from raw imaging data. T1-weighted magnetic resonance imaging (MRI) brain scans capture detailed structural architecture and may be used as a rich source of data for advanced machine learning models. Deep learning has been widely applied to medical imaging tasks such as disease prediction, detection and diagnosis, segmentation of abnormalities, and medical image synthesis^12^. Deep neural network models are capable of recognizing meaningful features and patterns from medical images that are difficult for clinicians to observe, leading to more accurate and efficient medical analyses^13^.

Previous works on brain imaging GWAS have primarily focused on image-derived phenotypes (IDPs) via image processing pipelines^14–16^. IDPs are generally used to capture high level features of brain anatomy, such as structural, functional, or connectivity patterns. These features are typically extracted through complex image processing steps to quantify specific aspects of brain structure or function. However, IDPs can miss subtle or complex patterns due to their reliance on predefined features and model assumptions, even more so in case of limited data availability. Additionally, variability in image quality and challenges in interpretation can impact the accuracy and applicability of these phenotypes across different populations or imaging modalities. IDPs often rely on segmentation as a critical step and in the case of brain imaging, this is difficult to achieve because of the brain’s complex anatomy, boundary ambiguity, and artifacts or noise induced from motion during the scanning process.

Recent advances in machine learning techniques offer more flexible methods that can learn important features from raw imaging data^17–21^. There are attempts using supervised deep learning to derive IDPs through labeled MRI data for brain imaging GWAS^22^. The primary challenge with this approach is that labeled data for supervised deep learning carry the same biases from the human labeler. Also, labeling is a labor intensive task, especially for large datasets.

Using unsupervised deep learning to discover phenotypes for GWAS of brain imaging can bypass the challenges of relying on predefined features, segmentation, and manual annotation by enabling models to learn complex and relevant features directly from the raw image data, where the model creates an *n*-dimensional representation known as Unsupervised Deep learning derived Imaging Phenotypes (UDIPs). GWAS on these UDIPs identified 97 unique loci^23^, many of them were missed by traditional IDP GWAS^14,16^.

Here, we propose an approach where an unsupervised learning vision transformer (ViT)^24^ model is used for brain imaging GWAS. This utilizes a different inductive bias from CNN architectures, which typically build representations through stacked layers of template matching, whereas the ViT uses patch-based self-attention to directly model potentially long-range interactions within the image. ViTs have demonstrated strong performance in various medical imaging applications, including feature extraction from brain MRI^25^, their architectural design facilitates the direct modeling of global context and long-range dependencies, even in the early layers^24,26^.

This paper presents a novel Vision Transformer autoencoder to derive a new class of phenotypes, ViT-UDIPs, directly from brain MRI data. A genome-wide association study performed on these ViT-UDIPs reveals novel genetic loci that were not captured by previous analyses. Additionally, an interpretability framework is introduced to probe the model’s learned pattern, offering insights into the ViT-derived phenotypes.

## Methods

Our overall pipeline is shown in Figure 1. The four major steps are data selection, ViT-AE model development, GWAS, and finally, SNP-ViT-UDIP to brain mapping via the PerDI approach.

**Figure 1.**
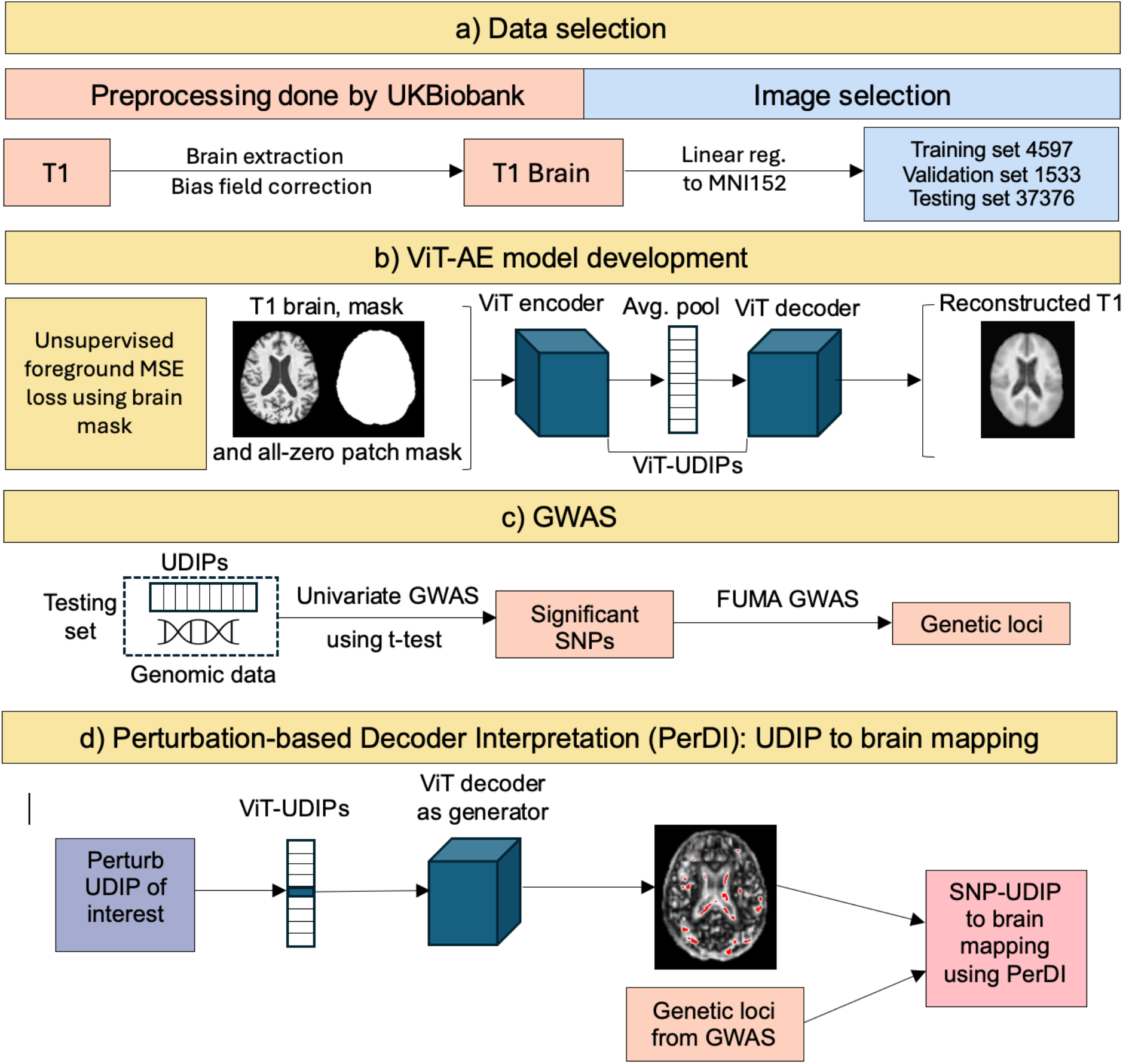
Overall pipeline of the study. a) T1 brain MRI preprocessed by UKBB were used for training, validation and testing. b) ViT-AE trained by background masked mean square error (MSE) loss of non-zero patches. c) Genetic loci discovered by univariate GWAS and FUMA GWAS. d) SNP-ViT-UDIP to brain mapping performed using Perturbation-based Decoder Interpretation (PerDI).

### Data Selection

The MRI scans were obtained from UKBB. The preprocessing done by UKBB involved brain extraction, bias field correction and registrations to align the imaging data into MNI152 space. While UKBB performed nonlinear registration, we applied only the affine component. Disjoint datasets were used for training, validation and GWAS to ensure robust model evaluation and prevent data leakage. 6,130 MRI scans were chosen for the model and split into 75/25 training/validation sets. 4,597 scans were used for training and 1,533 scans were used for validation. The dataset used for GWAS consisted of 37,376 scans.

The data-loading stage included pre-processing steps (see Supplementary Figure 1) on the linearly registered T1 brain MRI scans of dimensions 182 × 218 × 182. To allow division into equal-sized patches, these scans were padded to 182 × 224 × 182. The background voxels were already set to 0 by UK Biobank preprocessing. For the foreground voxels, we applied a z-transform to standardize the intensity values, ensuring consistency across scans and facilitating better model performance.

### ViT-AE Model

The ViT-AE architecture consisted of a ViT encoder, an average pooling layer and a ViT decoder. The features from the average pooling layer, which serve as a concise representation of the input, became a distinct token embedding that fed into the decoder layers along with empty token embeddings solely containing positional encoding. These combined token embeddings passed through multiple transformer layers and were decoded by a linear projection to reconstruct the input patches.

Figure 2 shows the block diagram of the overall ViT architecture. The T1 brain MRI images were segmented into non-overlapping patches of size 14 × 16 × 14 for a total of 13 × 14 × 13 patches. We optimized the model by dropping those patches which were entirely in the background across the whole batch^27^. Notably, this reduced the encoder workload by approximately 60%. To distinguish these masked patches from brain mask in our pipeline, we referred to them as *batch mask*. The hyperparameters used in the ViT-AE model are given in Table 1.

**Table 1.** ViT-AE hyperparameters.

| Parameter | Setting |
| --- | --- |
| Patch size | 14 x 16 x 14 |
| Total number of patches | 12 x 14 x 13 |
| Batch size | 2 |
| Encoder embedding dimension | 128 |
| Number of attention heads | 8 |
| Number of layers | 12 encoder, 12 decoder |
| Optimizer | AdamW |
| Learning rate | LR scheduler, lr=0.001, gamma=0.5, step size=3, minimum clipped at 1e-5 |
| Decoder embedding dimension | 64 |

**Figure 2.**
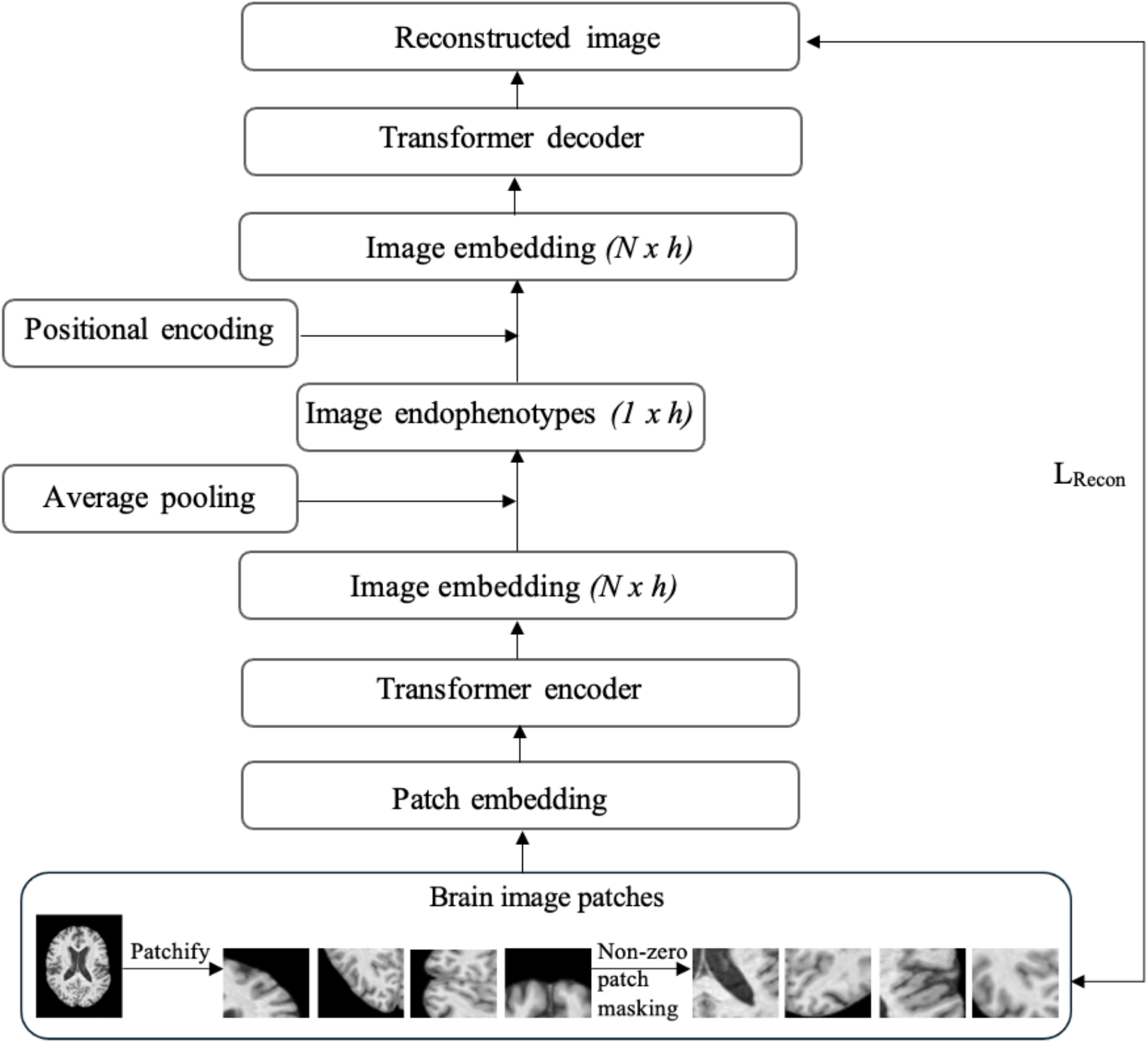
Block diagram of ViT-AE model, showing the generation of the endophenotypes from the average pooling layer which are then used by the ViT decoder for prediction of output patches. With *N* being number of tokens and *h* being hidden dimensions, the encoder creates image embeddings of size *N* × *h* which are averaged across *h* dimensions to create the ViT-UDIPs. Empty patch embeddings with positional encoding are used to re-create *N* × *h* image embeddings for the decoder. Reconstruction loss is denoted by *L*_*Recon*_.

MSE loss was computed between predicted patches and original patches within the batch mask. Figure 3 shows the ViT-AE modules and high-level pipeline of image and latent processing from encoder to decoder, as well as the final stage of loss computation. MSE loss was subsequently backpropagated to optimize model parameters during training.

**Figure 3.**
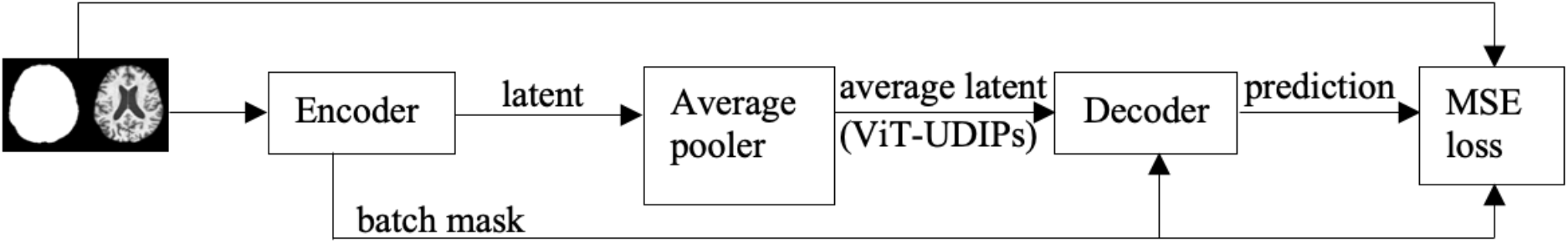
Image and latent flow through ViT-AE modules

The reconstructed layer assembles and reshapes the predicted patches from the decoder into 3D images of size 182 × 224 × 182, as shown in Figure 4.

**Figure 4.**
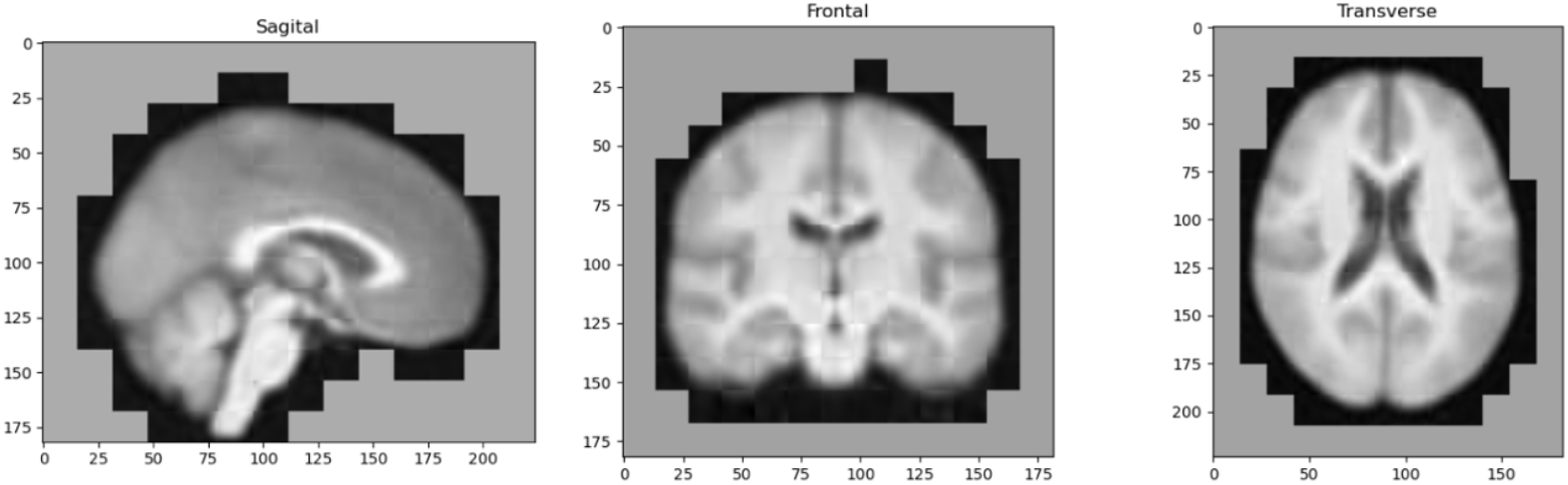
Reconstructed brain images with batch mask. Note that the gray areas in the images correspond to the patches which were masked due to containing all-zero values across all three image axes.

The ViT-AE model was trained for 400 epochs on training set and validated on a separate validation set. GWAS was later performed on the held-out test set using the ViT-UDIPs extracted by the trained model.

### GWAS

Related individuals were filtered out since our GWAS was conducted using a linear model, additionally, multiple visits were filtered where only scans from the first visits were retained. A total of 22,250 subjects were used for performing GWAS on the extracted ViT-UDIPs. GWAS was performed between the SNPs and the ViT-UDIPs, with age (UKBB field 21003), sex (UKBB field 31), 10 genetic principal components (UKBB field 22009), ethnicity (UKBB field 21000), T1 inverted contrast to noise ratio (UKBB field 25735), assessment center (UKBB field 54) as covariates, using a GPU-based GWAS tool developed in-house, where the p-value threshold was set to 5 × 10^−8^/128. We created a summary statistics file with only the most significant p-value (minP) for each SNP. This file was uploaded and run on FUMA SNP2GENE^28^ and the generated GenomeRiskLoci.txt file was then used to obtain the clumped loci.

The top five ViT-UDIPs with the highest number of significant SNPs were used for the next step, where noise was added to perturb these ViT-UDIPs for decoder interpretation.

### PerDI

To visualize the brain regions associated with the extracted ViT-UDIPs, we utilized a perturbation-based approach^23^. For 500 subjects from the test set, we added 1 standard deviation of noise to each of the five dimensions out of the 128 ViT-UDIPs, with the highest number of significant SNPs. The decoder was then used to reconstruct the images based on the perturbed ViT-UDIPs, followed by a voxel based paired t-test as shown in Figure 5.

**Figure 5.**
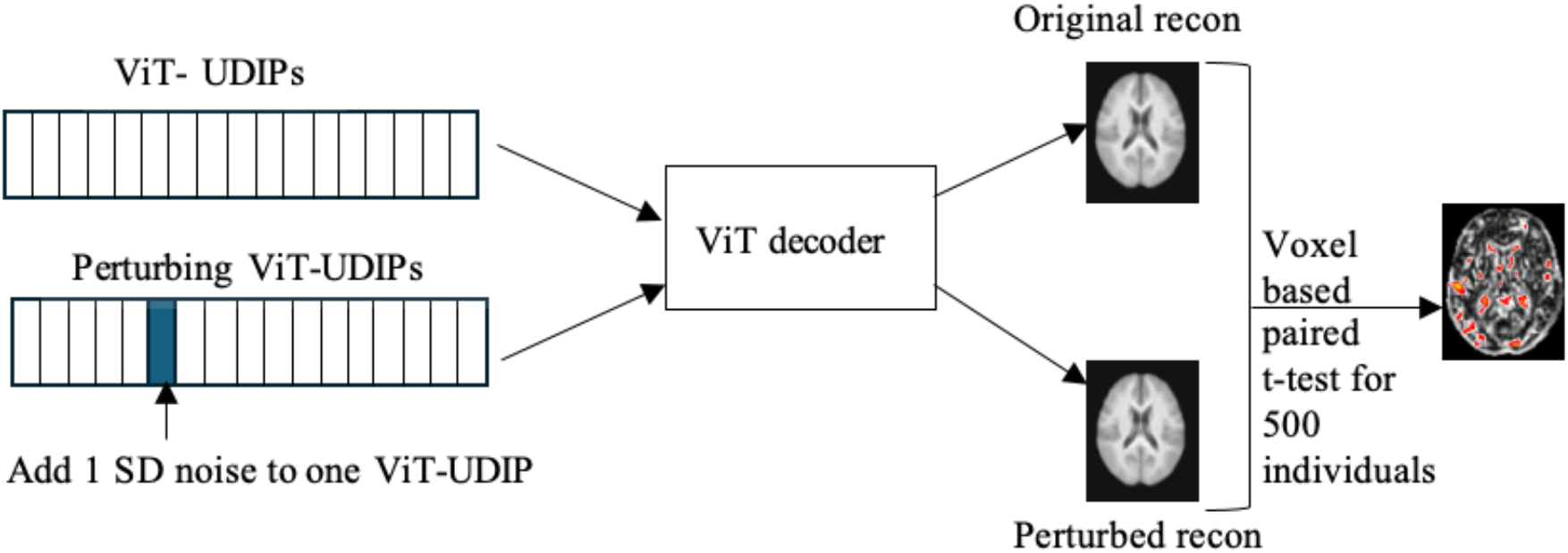
Perturbation-based Decoder Interpretation (PerDI)

A mask based on a brain template linearly registered to the T1 MNI152 space was applied to the t-test results, followed by a Gaussian filter (sigma=1) for smoothing. The five resulting images, one for each of the perturbed ViT-UDIPs, were saved as NIFTI files for visualization using FSLeyes. These NIFTI files were also used to find the regions of intersection with the Harvard-Oxford Cortical and Subcortical Structural Atlases (RRID:SCR_001476)^29–32^ and MNI Structural Atlas^33,34^.

## Results

### Model Performance

The average validation loss of the ViT-AE model remained at approximately 0.253 after 180 epochs, as shown in Figure 6. Average loss on test set was 0.2598. Training was stopped when we did not see any improvement in the number of discovered loci.

**Figure 6.**
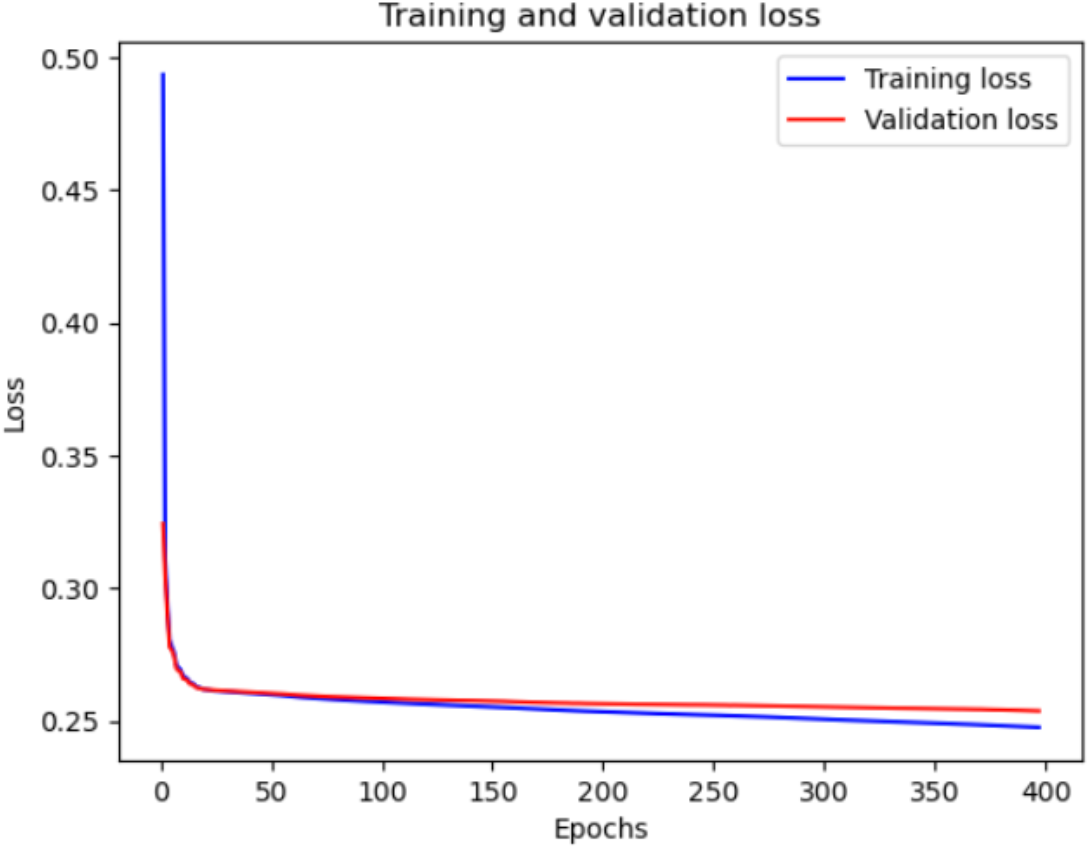
Training and validation loss of ViT-AE

Figure 7 displays brain images of two individuals, highlighting visible differences in anatomical structure, along with their respective reconstructed images by the ViT-AE model, where these differences appear to be preserved.

**Figure 7.**
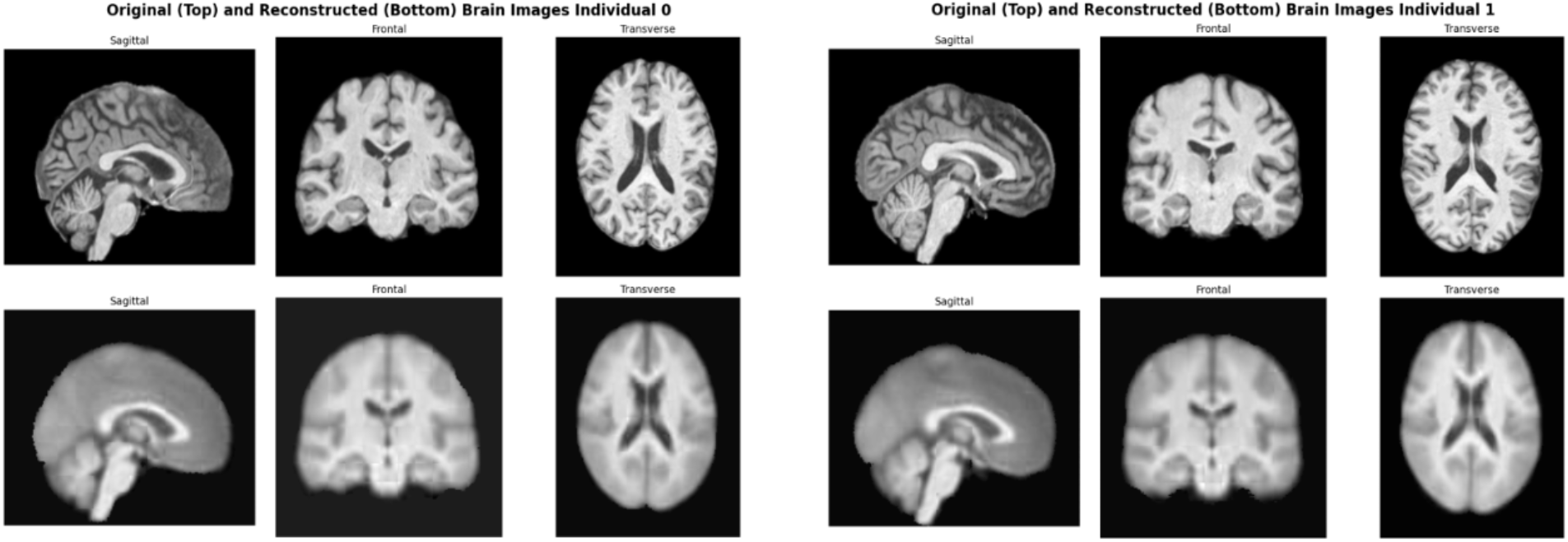
Original and reconstructed brain images for two individuals

As a comparison, the CNN-based UDIP model^23^ achieved an average validation loss of approximately 0.28 after 15 epochs that increased to 0.3 at 25 epochs (see Supplementary Figure 2). Visual observation of reconstruction was similar to that of the ViT-AE model (see Supplementary Figure 3).

Our ViT-AE model was developed for representation learning from brain MRI scans and extraction of ViT-UDIPs. Our workflow pipeline did not focus on hyperparameter tuning. However, initial implementation of the ViT-AE model consisted of encoder embedding dimension of 384 and this was later changed to 128 (see Supplementary Note 1 and Supplementary Figure 4A, 4B). The final pipeline included a step learning rate scheduler with a step of 3 and decay rate of 0.5 and AdamW optimizer with an initial learning rate of 0.01 (see Supplementary Note 2).

### Statistical Analysis of Reconstruction Loss

We performed basic statistical analysis of inferencing results to determine distribution of reconstruction loss and outliers in the dataset (see Supplementary Note 3). Although the primary goal of our work was to design a ViT-AE-based pipeline for brain feature extraction and finding associated genetic signals, this analysis provided insights into the model’s performance and helped detect anomalies and unusual deviations that could indicate potential challenges in reconstruction. Overall reconstruction loss was found to be distributed normally, suggesting the model performed consistently across most scans in the dataset (see Supplementary Figure 5A). Boxplot and scatter plots indicated outliers that might have contributed to higher average reconstruction loss for some scans (see Supplementary Figure 5B, 5C). For the sake of efficiency, we considered only extreme outliers, which we defined to be outside of lower quartile (Q1) and upper quartile (Q3) by three times the inter-quartile range (IQR) (see Supplementary Figure 5D) and visualized those with the highest reconstruction losses to determine if there were significant anomalies in the brain structures or artefacts introduced by motion during scanning (see Supplementary Figure 6).

Analysis of ViT-UDIPs obtained from inferencing demonstrated normalized distribution for each ViT-UDIP (see Supplementary Figure 7). Lines in violin plot for the ViT-UDIPs represented median, Q1 and Q3 values and each violin width represented kernel density estimate (KDE) of the data. The overall shapes indicated overall normality of the ViT-UDIPs.

### GWAS

GWAS was done on the test dataset using the features from the checkpoints saved at every fixed number of epochs. We conducted GWAS using linear model to determine the p-value for each SNP-ViT-UDIP pair, then used FUMA to clump the significant SNPs (those with the minimum p-value passing the Bonferroni-corrected threshold) into loci. We then identify which model(s) yielded the highest number of loci associated with brain structure. During training, we recorded the number of loci discovered every 10 epochs. The model was trained until 100 consecutive epochs showed no further increase in the number of loci identified. The highest number of loci was discovered using models trained for 200, 220 and 300 epochs (see Supplementary Figure 8).

We only analyzed FUMA GWAS results for epochs 200, 220 and 300, as they represent the epochs with the most number of loci discovered. Notably, we discovered 10 new loci associated with brain structure that were not previously reported by CNN-based UDIP approach^23^. Using dbSNP^35^, we investigated the lead SNPs in these loci. Figure 8 shows the previously unreported loci and the corresponding dimension locations of UDIPs on a Manhattan plot. Chromosomes 7 and 11 each had two significantly associated loci. An example of how to interpret the plot is as follows: the gene SLC6A20 was identified as significantly associated with the ViT-UDIP located on dimension 94, indicating a relationship between the genetic expression of SLC6A20 and the feature represented in dimension 94 of the learned embeddings.

**Figure 8.**
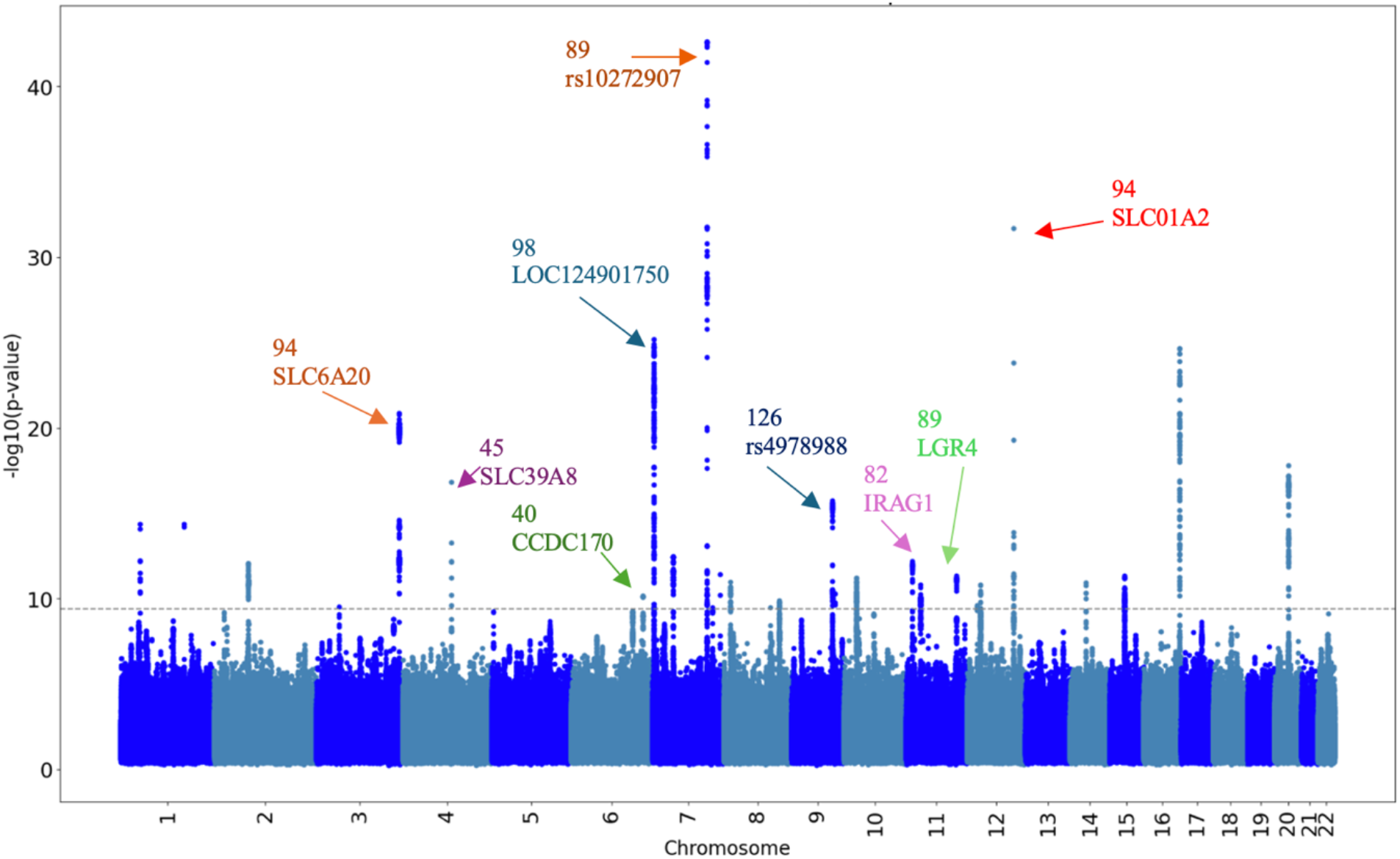
UDIP dimension location and new loci discovered from ViT-based UDIP approach

GWAS revealed a total of 29 loci (see Supplementary Table 1 for all loci and associated lead SNPs and genes^36^), out of which 10 loci were not previously reported in the CNN-based UDIP approach^23^. These were cross-checked with the GWAS catalog and our findings are shown in Table 2.

**Table 2.**
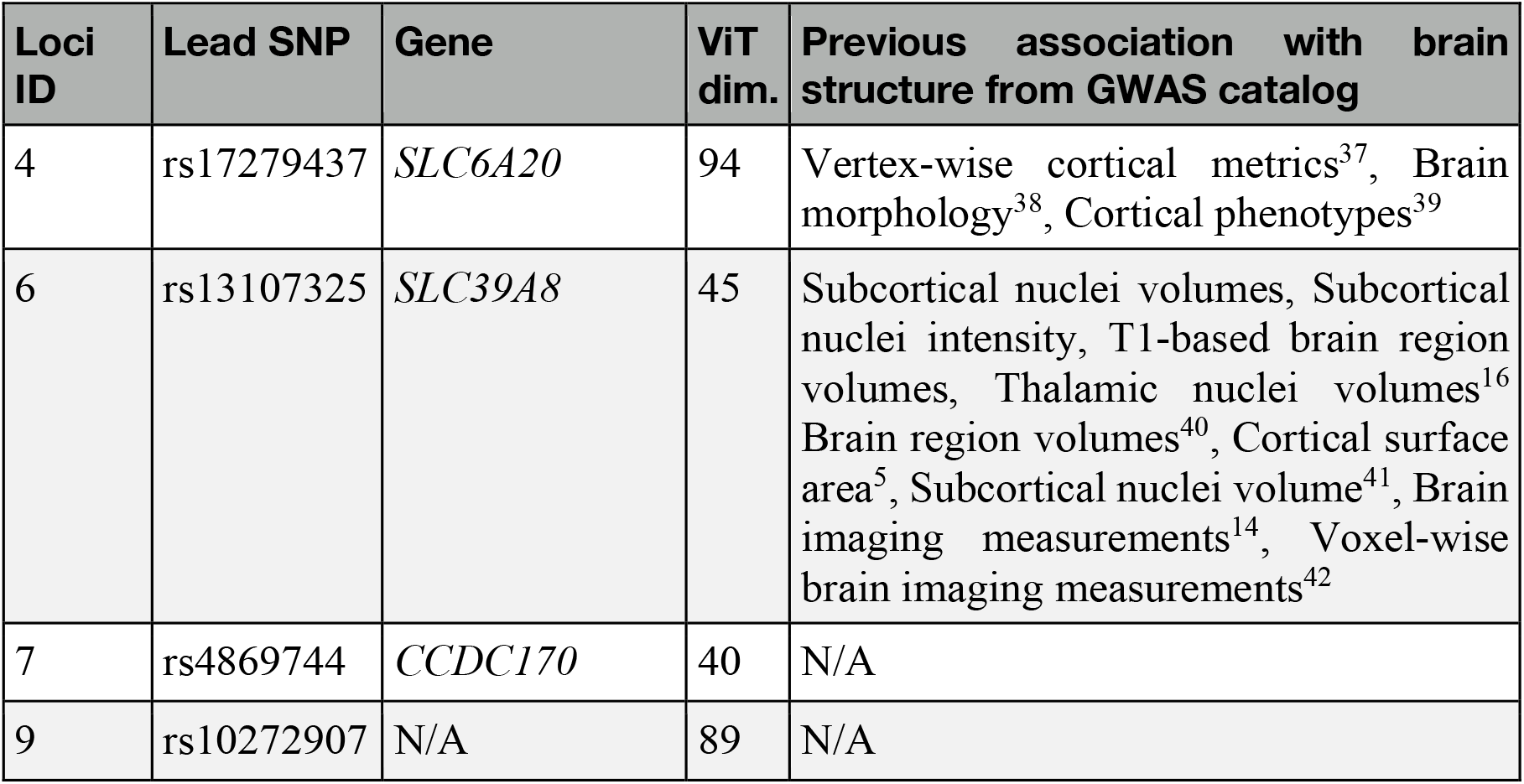
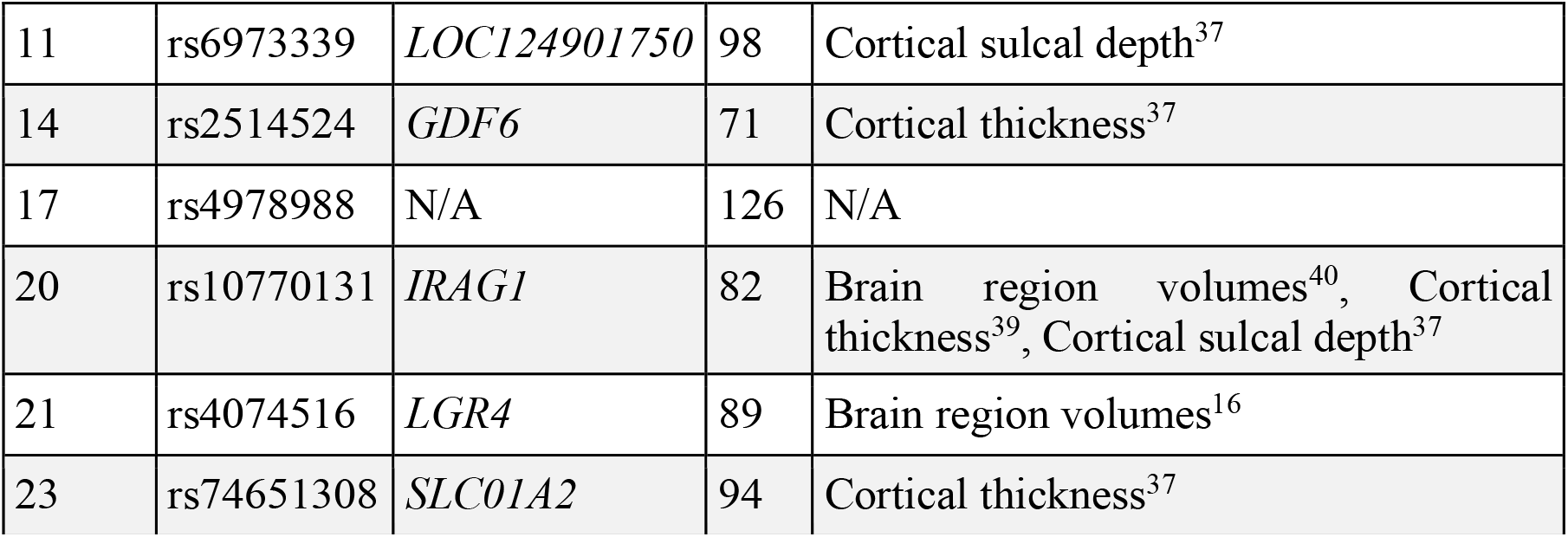
Loci cross-checked with GWAS catalog.

| Loci ID | Lead SNP | Gene | ViT dim. | Previous association with brain structure from GWAS catalog |
| --- | --- | --- | --- | --- |
| 4 | rs17279437 | *SLC6A20* | 94 | Vertex-wise cortical metrics<sup>37</sup>, Brain morphology<sup>38</sup>, Cortical phenotypes<sup>39</sup> |
| 6 | rs13107325 | *SLC39A8* | 45 | Subcortical nuclei volumes, Subcortical nuclei intensity, T1-based brain region volumes, Thalamic nuclei volumes<sup>16</sup> Brain region volumes<sup>40</sup>, Cortical surface area<sup>5</sup>, Subcortical nuclei volume<sup>41</sup>, Brain imaging measurements<sup>14</sup>, Voxel-wise brain imaging measurements<sup>42</sup> |
| 7 | rs4869744 | *CCDC170* | 40 | N/A |
| 9 | rs10272907 | N/A | 89 | N/A |
| 11 | rs6973339 | *LOC124901750* | 98 | Cortical sulcal depth<sup>37</sup> |
| 14 | rs2514524 | *GDF6* | 71 | Cortical thickness<sup>37</sup> |
| 17 | rs4978988 | N/A | 126 | N/A |
| 20 | rs10770131 | *IRAG1* | 82 | Brain region volumes<sup>40</sup>, Cortical thickness<sup>39</sup>, Cortical sulcal depth<sup>37</sup> |
| 21 | rs4074516 | *LGR4* | 89 | Brain region volumes<sup>16</sup> |
| 23 | rs74651308 | *SLC01A2* | 94 | Cortical thickness<sup>37</sup> |

Several loci had been previously associated with brain structure. For brevity, we included only those loci which were discovered by ViT-AE, but not by CNN-based UDIP approach. Furthermore, we limited our findings to traits derived from T1-weighted MRI only. Investigating the above loci, we found that loci 4, 6, 11, 14, 20, 21 and 23 were already present in the GWAS catalog.

Loci 7, 9 and 17 had not been previously documented in the GWAS catalog as being associated with brain architecture. Locus 7, characterized by the lead SNP rs4869744 and linked to the gene *CCDC170* on chromosome 6, was identified through ViT-AE dimension 40. Prior research had primarily associated this locus with bone density. Similarly, locus 9, marked by the lead SNP rs10272907 on chromosome 7, had also shown connections predominantly related to bone density. Locus 17 with lead SNP rs4978988 on chromosome 9 had previously been found related to increased signal intensity in white matter regions, typically observed in T2-weighted MRI or FLAIR sequences^43^. However, brain phenotypes derived from T1-weighted MRI had not been associated with this locus.

### Visualizing endophenotype activation patterns by PerDI

We performed PerDI on dimensions that a) included the highest number of SNPs significantly associated with the ViT-UDIPs (dimensions 18, 34, 40, 124, 126), and b) revealed new loci previously not discovered by CNN-based UDIP approach (dimensions 45, 82, 89, 94, 98).

Using FSLeyes, we visualized NIFTI files for each of these dimensions. We identified activated regions by overlaying the images with the Harvard-Oxford Cortical and Subcortical, and MNI Structural Atlases.

Applying PerDI to ViT-UDIPs revealed learned representations from brain MRI scans. Left-right symmetry was observed in these visualizations. Figure 9 demonstrates the activation regions for dimension 98. At the subcortical level, the thalamus region was seen highlighted on both right and left sides of the brain, along with cerebral white matter and cerebral cortex. The cerebellum, along with the frontal and parietal lobes, was highlighted, capturing a range of structural patterns across the hindbrain and forebrain. Similar patterns were identified across the other dimensions, highlighting the model’s ability to generalize its representations effectively.

**Figure 9.**
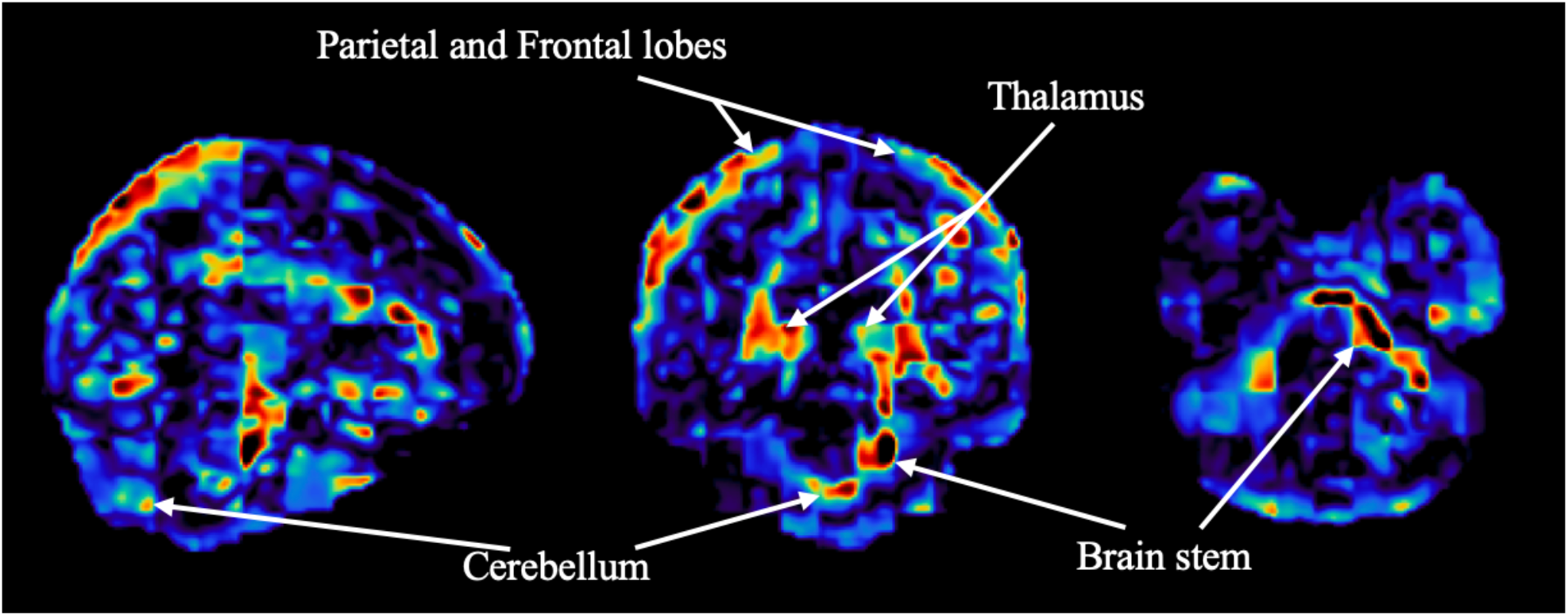
PerDI activation on dimension 98

PerDI activation regions were investigated for loci 7, 9 and 17, which were previously unreported to have associations with brain structure. These associations were identified by dimensions 40, 89 and 126, respectively. Figure 10 provides the activations regions from each of these dimensions.

**Figure 10.**
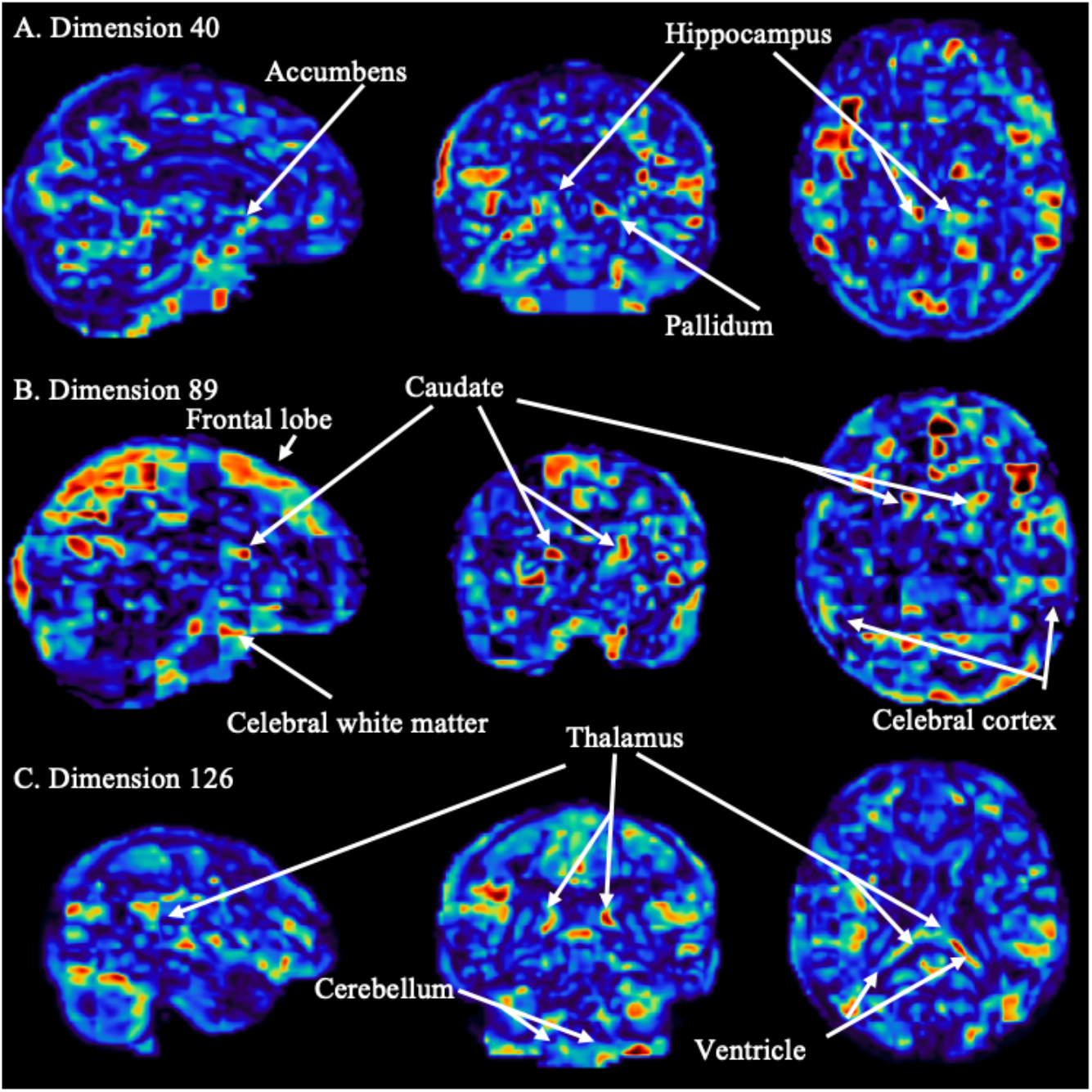
Activation regions for dimensions 40, 89, 126

Dimension 40 (Figure 10A), visualized using PerDI, highlights the pallidum, hippocampus, and accumbens in a symmetrical pattern. The frontal lobe region, as well as the caudate in the central subcortical region of the brain is highlighted by dimension 89 (Figure 10B). In a similar way, dimension 126 (Figure 10C) activates the ventricles and thalamus in the central subcortical region. We also see activation in the cerebellum. In all three dimensions, we also see activation in regions corresponding to the subcortical regions of cerebral cortex and cerebral white matter.

The PerDI ViT-UDIP SNP-to-brain mapping demonstrated the ViT-AE model’s ability to capture activations across the brain, revealing structural patterns at varying levels of detail. Specific regions such as the pallidum, hippocampus, accumbens, thalamus, and the cerebral cortex were highlighted, showing activation within central subcortical and cortical areas. Additionally, activations were observed in key structures such as the ventricles and cerebral white matter, which play crucial roles in brain communication and the protection of neural tissue. These activations reflected the model’s capacity to identify symmetrical patterns across bilateral structures, consistent with the brain’s inherent anatomical symmetry, and to recognize fine-grained structural details.

### Attention Head Analysis

We visualized the attention patches on brain atlases using FSLeyes. Our analysis provided insights into the model’s interpretability, highlighting the regions of the brain that are most influential in the feature extraction process and offering a deeper understanding of how the ViT-AE model captures complex spatial dependencies in brain MRI scans.

Our observations revealed long-range dependencies and symmetrical attention across the brain regions. The model was able to attend to distant patches across different brain regions simultaneously, demonstrating its capacity to capture non-local relationships within the brain (see Supplementary Figure 9A). Additionally, we observed symmetrical attention, with patches in one hemisphere of the brain attending to corresponding patches in the opposite hemisphere (see Supplementary Figure 9B). This symmetry suggests that the ViT-AE model is effectively capturing bilateral relationships, reflecting the natural symmetry of brain anatomy. These findings highlight the model’s ability to integrate both local and non-local features, providing a comprehensive understanding of brain architecture and uncovering complex interconnections between brain regions.

Furthermore, we investigated the attention scores of the trained model and found some interesting patterns. From the very first attention layer, the model started to focus on a subset of patches across different heads. The attention pattern of each head was extremely sparse (Figure 11), while different heads at different layers focused on different patches. This subset of patches consisted of less than one-third of the total patches, with all other patches attending to them.

**Figure 11.**
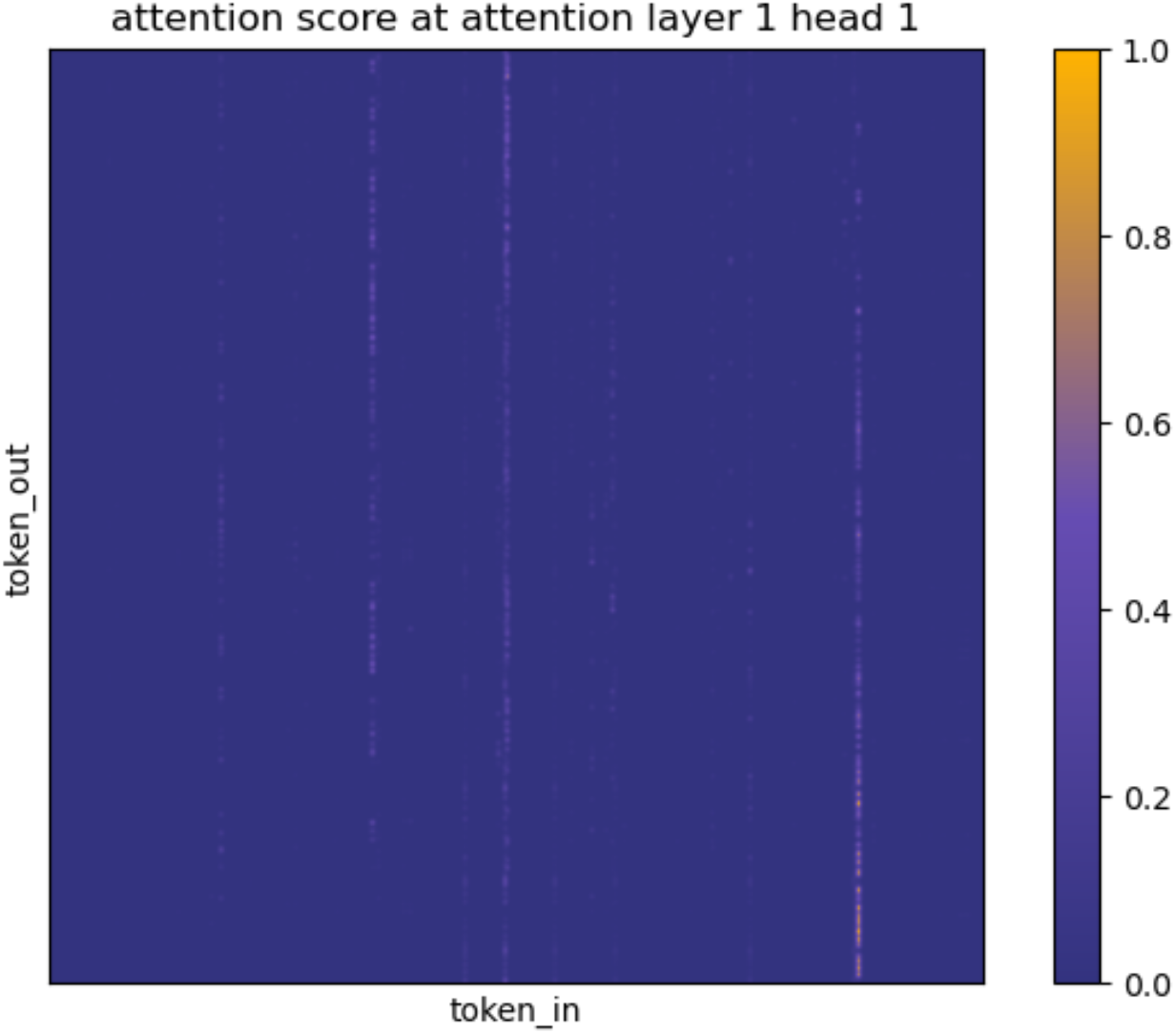
Softmax attention score at attention layer 1 head 1 between 957 tokens, image is blurred using Gaussian filter with sigma=2 for better visual quality.

We found that this behavior is driven by the specific embeddings learned by the model. The input embedding matrix projects the majority of patches to a similar direction, while a small subset remains weakly correlated with each other and the majority (Figure 12). Henceforth, we will refer to them as audience tokens and spotlight tokens for brevity.

**Figure 12.**
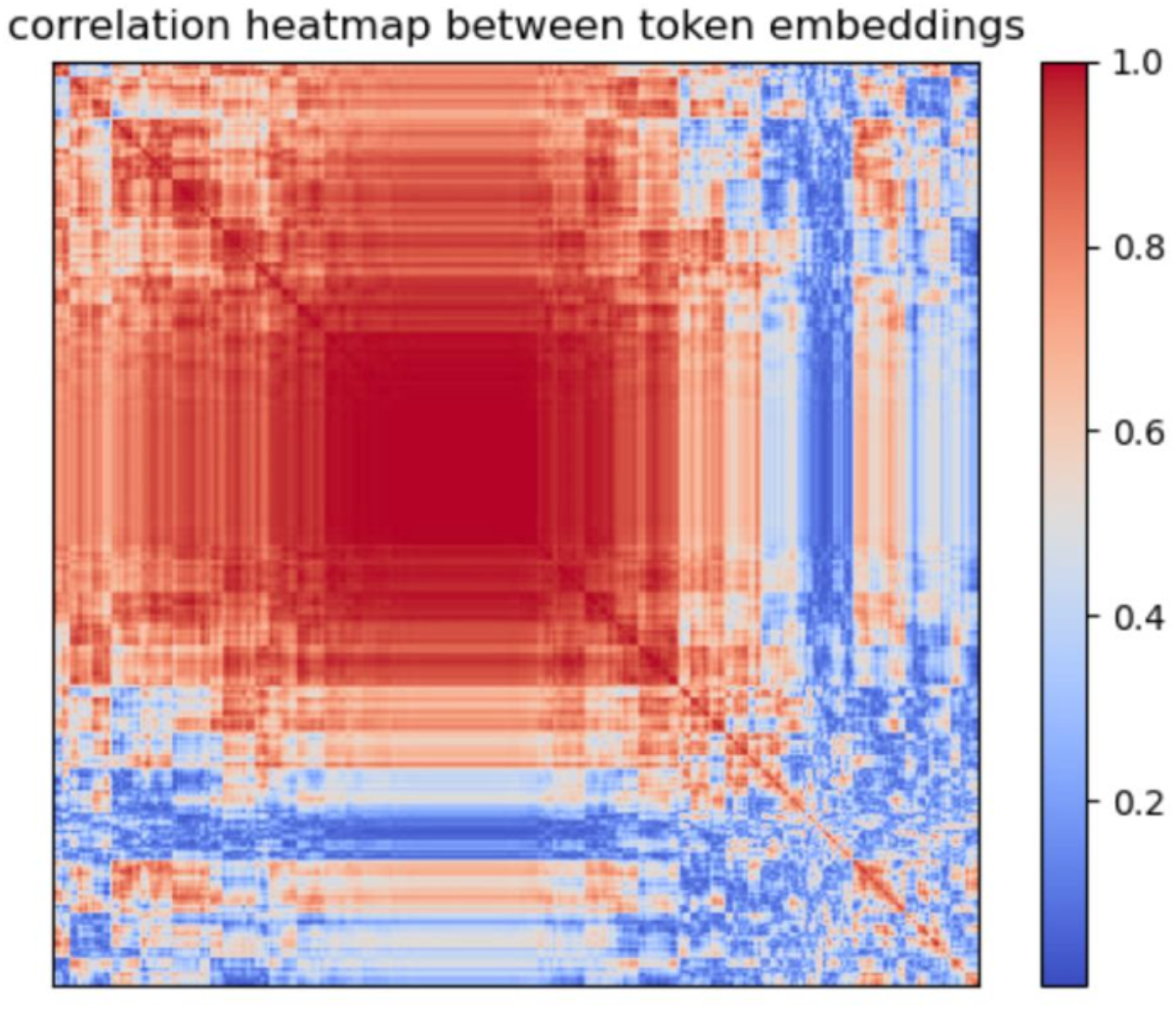
Correlation heatmap between 957 input embeddings, we use hierarchical clustering with average linkage to reorder the dimensions of the heatmap.

At each attention head, the (unnormalized) attention score between two tokens *X*_1_ and *X*_2_ can be written as 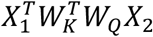,this can be interpreted as first rotating them differently (based on the rotation matrices formed by the left and right singular vectors of 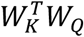), followed by projecting the rotated *X*_1_ and *X*_2_ on to the dimensions with nonzero singular values and finally computing a weighted inner product. Specifically, singular value decomposition of 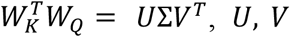 are rotation matrices and Σ is a diagonal scaling matrix. The interaction between *X*_1_ and *X*_2_ can be written as (*PU*^*T*^*X*_1_)^*T*^Σ_*r*_(*PV*^*T*^*X*_2_), where *P* is a wide identity matrix that selects the first *d*_*h*_ coordinates of the input, the dimension of the head. Σ_*r*_ is the reduced singular value matrix which only consists of nonzero entries of Σ of shape *d*_*h*_ × *d*_*h*_, As a result, at each attention head, audience tokens tend to focus on one or a few nearby spotlight tokens that happen to align with them through rotations, leading to sparse attention patterns and accumulation of information from these spotlight patches.

We then visualize the distribution of spotlight tokens across different brain slices. The highlighted regions in Figure 13 indicate key locations where the model focuses its attention. We observe that these spotlight patches tend to cluster around deep sulci, align along the boundaries of the ventricles, and appear in structures such as the cerebellum and brainstem. Additionally, some spotlight patches appear in corresponding regions across hemispheres, suggesting a tendency to capture bilaterally relevant anatomical features. The distribution of attention is sparse and concentrated around major structural landmarks, indicating that the model prioritizes these regions for information aggregation.

**Figure 13.**
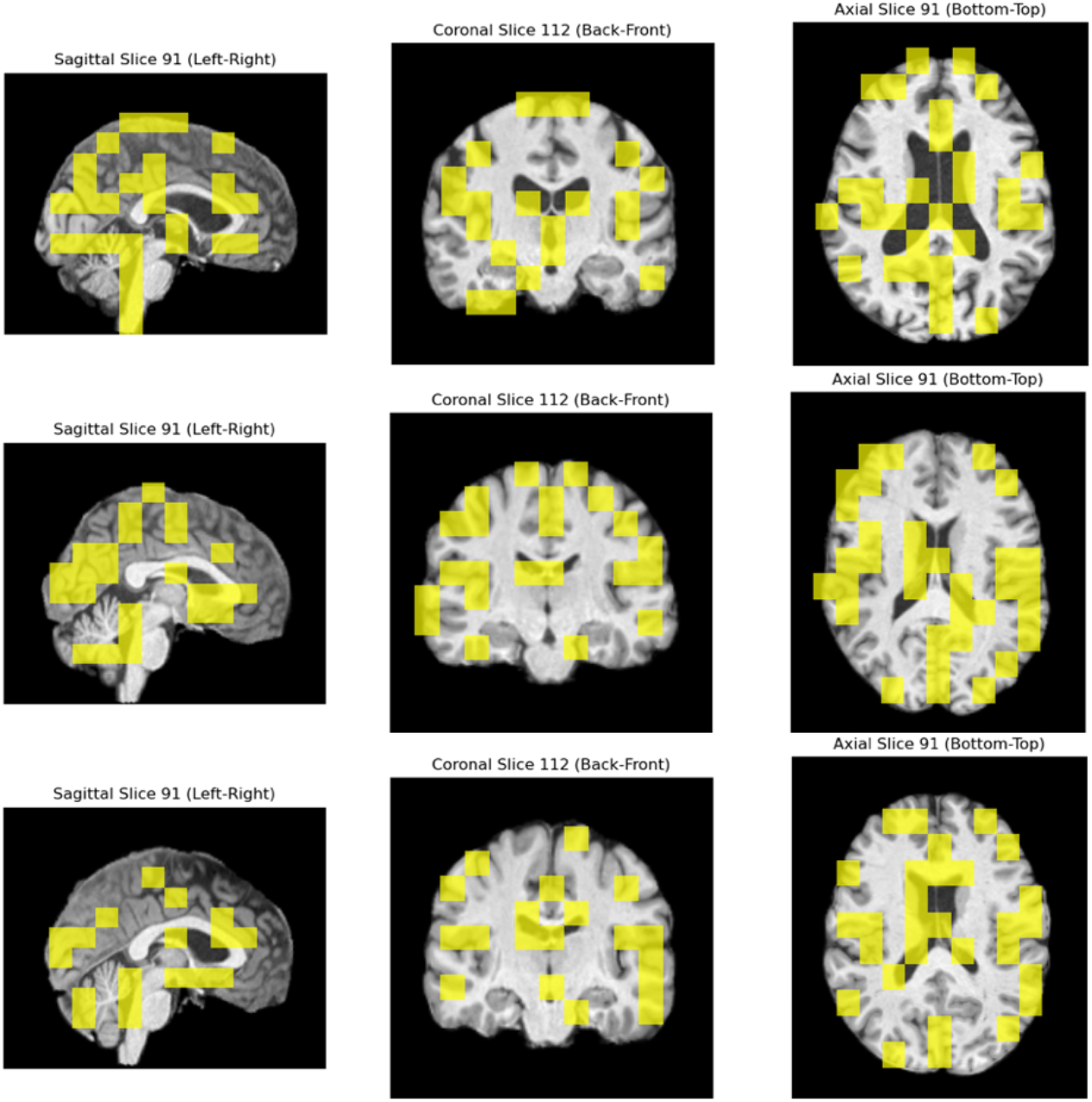
Visualization of spotlight tokens across brain slices. Highlighted regions (yellow)

## Discussion

A central theme of our study is that a different architectural inductive bias can reveal new biological insights from the same data. We developed an approach that integrated unsupervised deep representation learning, specifically using a ViT-AE model, for brain imaging GWAS. Our ViT-AE pipeline successfully identified 10 genetic loci associated with brain structure that were missed by a previous CNN-based approach, with three of these (loci 7, 9, and 17) having no prior associations with brain morphology in the GWAS Catalog. We attribute this success to the Vision Transformer’s inherent ability to model flexible dependencies through its self-attention mechanism. Unlike CNNs, which build a hierarchy of local features, the ViT can directly capture long-range patterns, such as the left-right hemispheric symmetry and relationships between distant sulci. This suggests that the genetic influence of these novel loci may manifest as subtle, distributed, or symmetrical patterns across the brain.

The model’s ability to uncover these novel associations stems from its unique architectural properties. Our pipeline also incorporated a perturbation-based decoder interpretation approach to map these loci and their associated imaging phenotypes to specific brain regions. By comparing original and reconstructed images for two individuals and visualizing the absolute differences (see Supplementary Figure 10), we showed our model’s capacity to capture structural differences in brain morphology. Furthermore, our analysis of the model’s attention mechanism revealed the data-driven emergence of “spotlight tokens,” which receive a disproportionate amount of attention and correspond to anatomically informative regions with high structural variance, such as the boundaries between different tissue types (e.g., gray and white matter), the edges of ventricles, or the complex folding of deep sulci. The model appears to have learned to focus its attention on these information-rich patches.

Despite these promising results, our study has limitations that present clear avenues for future work. A key topic in current machine learning research is the performance of Vision Transformers (ViTs), particularly when faced with limited data. This is especially a concern in healthcare because of challenges in data availability^37^. As an experimental approach, we trained our ViT-AE model using the combined original training and testing datasets and validated it on a disjoint validation set. The model’s performance considerably improved with an average validation loss of 0.18 after 300 epochs, compared to 0.253 on the training set only (see Supplementary Figure 11A). The number of loci discovered was also significantly higher, with 96 associated loci found at 221 epochs (see Supplementary Figure 11B). Although the trained model identified additional associated loci, these findings were excluded from the main results because the GWAS analysis was conducted on the original testing dataset, which was included in the training process. This overlap posed a risk of data leakage, potentially exaggerating the model’s performance and compromising the validity of the results.

Nonetheless, we present these findings to illustrate the ViT-AE model’s potential to perform better with a larger training dataset. To address the data-hungry nature of ViTs, self-supervised algorithms such as masked autoencoders (MAEs) have been shown to mitigate this issue^44^. While this approach was not incorporated into our current pipeline, we will consider it as a promising future direction. In addition, we will consider data augmentation and synthetic data generation techniques. Data augmentation methods, such as random rotations, flips, and intensity variations, could be employed to create more diverse variations of the original brain MRI scans. Additionally, utilizing synthetic data generation techniques could allow for the creation of realistic, high-quality brain images, addressing challenges related to limited data availability.

Besides the challenge of data size, this study didn’t extensively optimize for the network architecture. Specifically, the pooling layer in our ViT-AE architecture averaged the latent embeddings across all patches, which may lead to diminished local information. Some modifications that we would like to explore further include using adaptive average pooling, where some spatial information could be retained. Notably, we tried to use attentive pooling or learned queries to pool the embeddings into multiple representations; however, the model experienced dimensional collapse, resulting in significantly worse performance compared to using average pooling to extract the embedding.

The observation of “spotlight tokens” also has direct implications for the choice of patch size. While our chosen patch size (14×16×14) proved effective, a smaller patch size could potentially enable the model to resolve even finer anatomical details, allowing spotlight tokens to pinpoint more granular features. This might be crucial for phenotypes linked to subtle variations in cortical thickness or the integrity of smaller structures. However, smaller patches lead to a much longer sequence of tokens, increasing computational cost and potentially making it harder for the model to train. The optimal patch size should therefore find a balance between feature resolution and the ability to model global context, we leave this for future study.

## Data Availability

All code, model checkpoints and results used and produced in the present study will be made available in the coming days.

## Ethics oversight

Our analysis was approved by UTHealth Houston committee for the protection of human subjects under No. HSC-SBMI-20-1323. UKBB has secured informed consent from the participants in the use of their data for approved research projects. UKBB data was accessed via approved project 24247.

## Acknowledgements

This work was supported by grants from the National Institute on Aging U01AG070112 and R01AG081398.

