## Supplemental Materials for "Vision Transformer Autoencoders for Unsupervised Representation Learning: Revealing Novel Genetic Associations through Learned Sparse Attention Patterns"

1. Supplementary Notes
2. Supplementary Figures
3. Supplementary Tables

### Supplementary Notes

#### Supplementary Note 1: ViT dimensions

Our initial ViT-AE model had 384 encoder embedding dimensions, resulting in 384 ViT-UDIPs. We performed statistical analysis on these extracted features to find their cross correlation. Strong cross-correlation of features (correlation coefficient > 0.8) indicated embeddings collapsing, revealing that the model's latent space was not well-separated, likely causing poor generalization to unseen data. Reducing the dimensionality of the model by adjusting the model's architecture to 128 embeddings allowed better feature separation, where the model was able to distinguish between different aspects of the data more effectively, thus minimizing overfitting.

#### Supplementary Note 2: ViT-AE training learning rate and optimizer

We used StepLR from the PyTorch package optim.lr_scheduler to decay the learning rate every *n* epochs. The parameters we used were:

Optimizer: AdamW
Starting learning rate: 1e-2
Gamma: 0.5
Step size: 3
Lower cap of learning rate: 1e-5

Learning rate started at 1e-2 with a lower cap set at 1e-5. The parameters gamma and step_size control the multiplicative factor of learning rate decay and period of learning rate decay, respectively. The learning rate scheduler was applied to an AdamW optimizer. A modified version of the ViT-AE model used Stochastic Gradient Descent (SGD) optimizer. However, there was no improvement in model performance. Therefore, AdamW was used in the final pipeline.

#### Supplementary Note 3: Statistical analysis of reconstruction loss

Our ViT-AE model was trained on 4,597 T1-weighted brain MRI scans, while the validation set was of length 1,533. Inference and subsequent GWAS were performed on 37,376 scans. The average validation loss was 0.253, while the average test loss was 0.2597. We performed basic statistical analysis of inferencing results to determine distribution of reconstruction loss and outliers in the dataset to gain insights into the model's performance.

We found 191 number of outliers outside of lower quartile (Q1) and upper quartile (Q3) by 1.5 times of interquartile range (IQR) and 14 number of extreme outliers which were 3 times IQR. Investigating the top 3 outliers, we identified very noticeable brain structure anomalies either because of brain atrophy or poor brain scan quality which may have been brought about by movement during scanning.

### Supplementary Figures

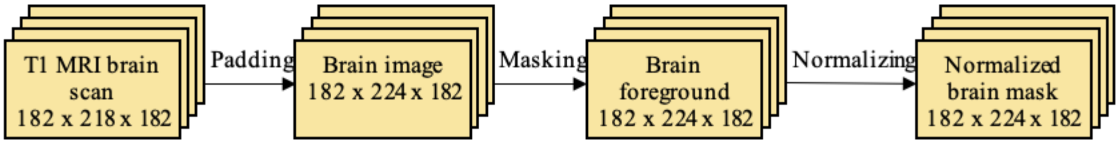


**Supplementary Figure 1 | T1 brain MRI preprocessing.** Pipeline shows pre-processing steps in dataloader. Original linearly registered T1 images were padded so that they could be divided into patches. Brain masking enabled brain region isolation, which were then normalized using z-transform to ensure consistency in intensity values.

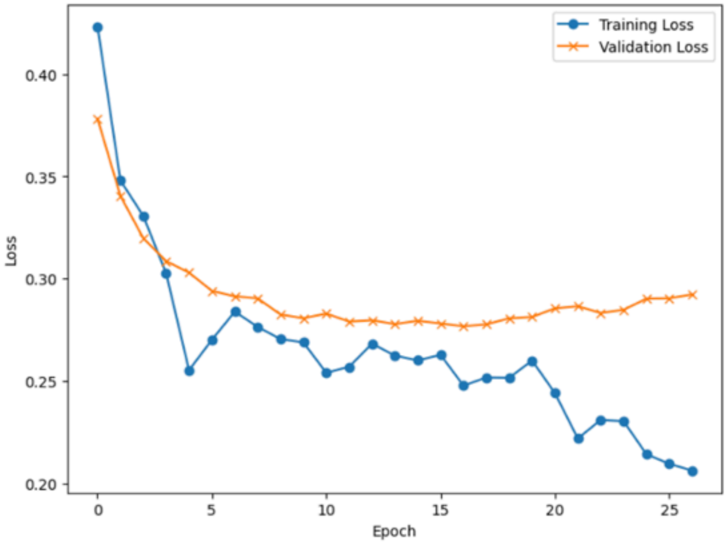


**Supplementary Figure 2 | Training and validation loss of CNN model for UDIP-based GWAS.** Average validation loss was 0.28 after approximately 15 epochs, before exhibiting overfitting.

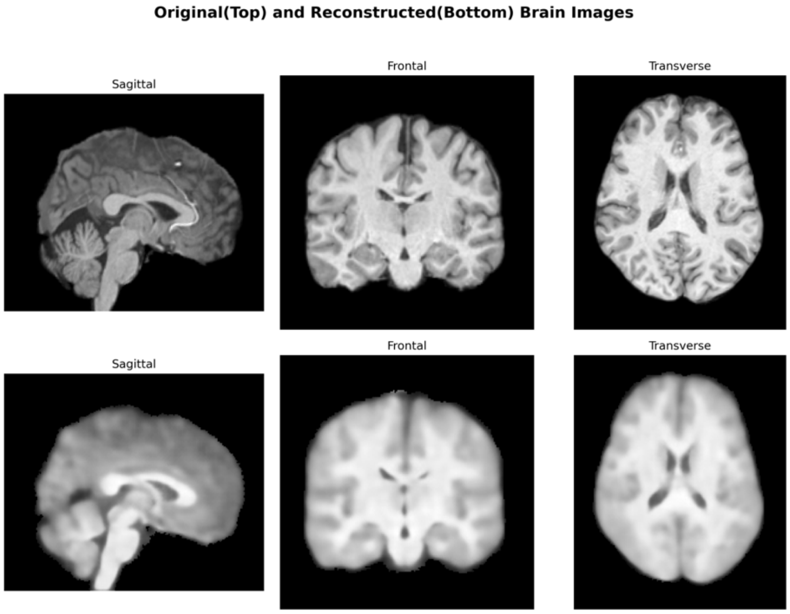


**Supplementary Figure 3 | Original and reconstructed brain images from CNN model for UDIP GWAS.** Visual observation of reconstructed images show similarity to ViT-AE model.

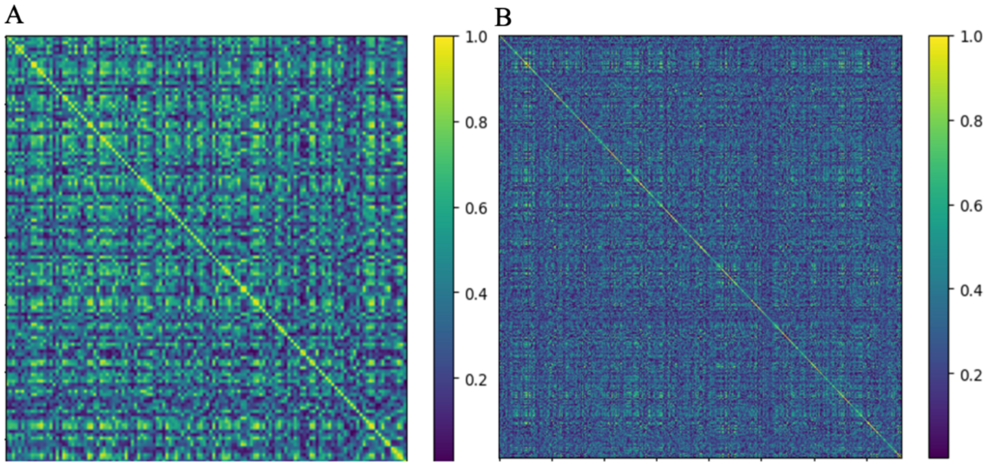


**Supplementary Figure 4 | Correlation coefficients of ViT-AE embeddings showing feature collapse in higher-dimensional ViT-AE. A.** The 384-dimension ViT-AE had strong correlation among embeddings that were possibly leading to over generalization of representations, thus resulting in poor model performance. **B.** The 128-dimension ViT-AE demonstrated less correlation resulted in better feature separation, allowing the model to distinguish between different aspects of the data more effectively, thus reducing the risk of overfitting.

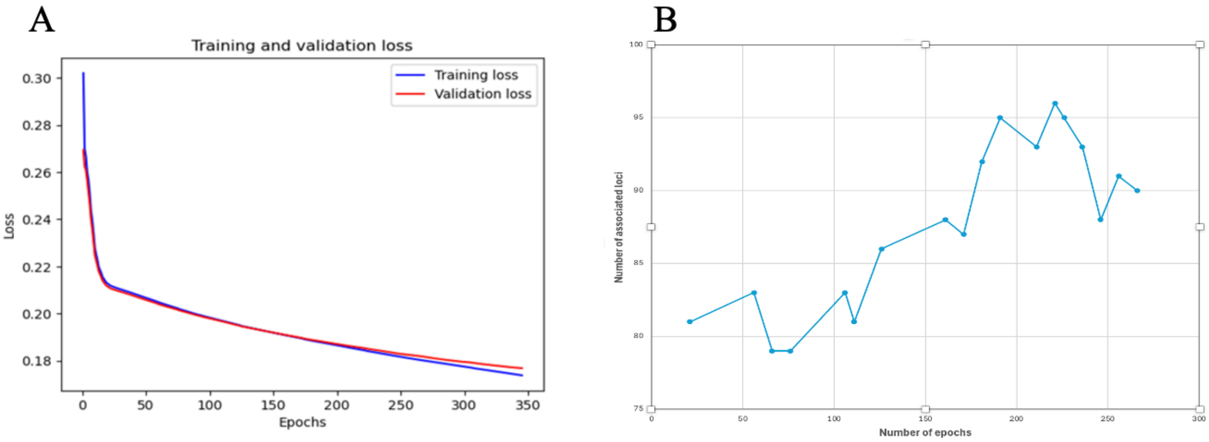


**Supplementary Figure 5 | Reconstruction performance and loci discovery of ViT-AE model trained on combined training and testing datasets. A.** Validation set remained disjoint from the combined datasets the model was trained on. However, average validation loss showed significant improvement compared to the model trained solely on the testing data. This demonstrated that a larger dataset would allow the ViT-AE to leverage its capacity more effectively, resulting in improved performance. **B.** Additionally, more associated loci were discovered by this trained model. However, these results were not included in our final pipeline because the GWAS was performed on the testing dataset, which was also used to train the model. This overlap could have led to data leakage, potentially inflating the model's performance and undermining the validity of the findings.

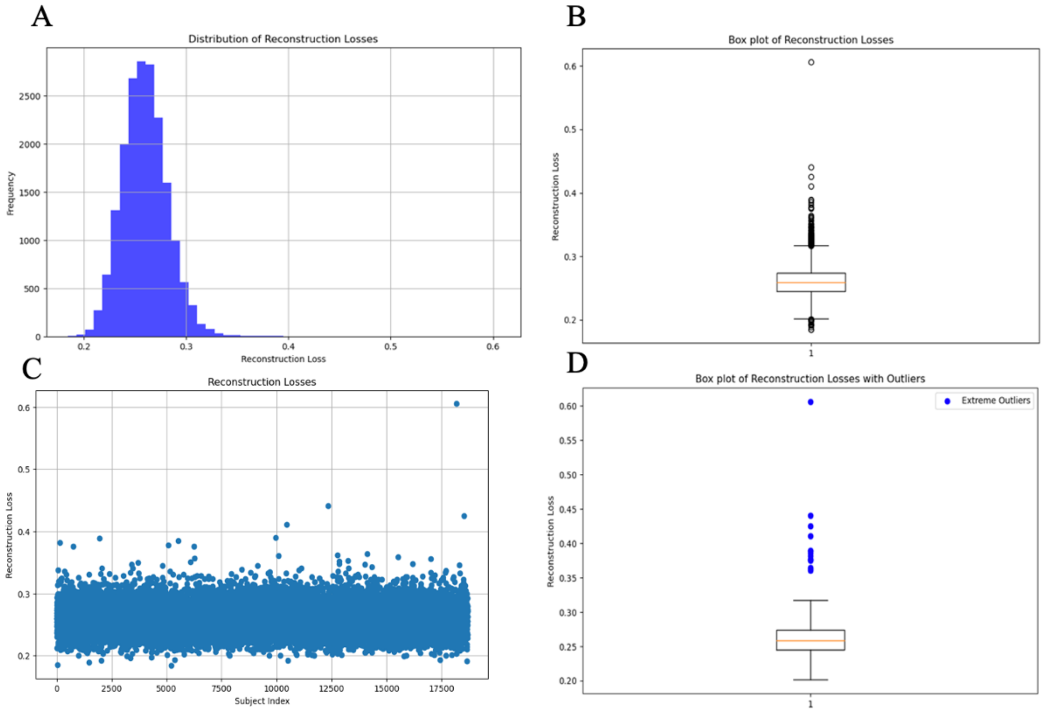


**Supplementary Figure 6 | Statistical analysis of inferencing results. A.** Overall distribution shows consistency in reconstruction loss for most images. **B.** Box plot shows outliers for reconstruction loss which are outside of Q1 and Q3 by 1.5 times of IQR. **C.** Scatter plot shows batch-wise reconstruction loss variation among 37,376 scans with batch size of 2. **D.** Box plot shows extreme outliers for reconstruction loss which are outside of Q1 and Q3 by 3 times of IQR.

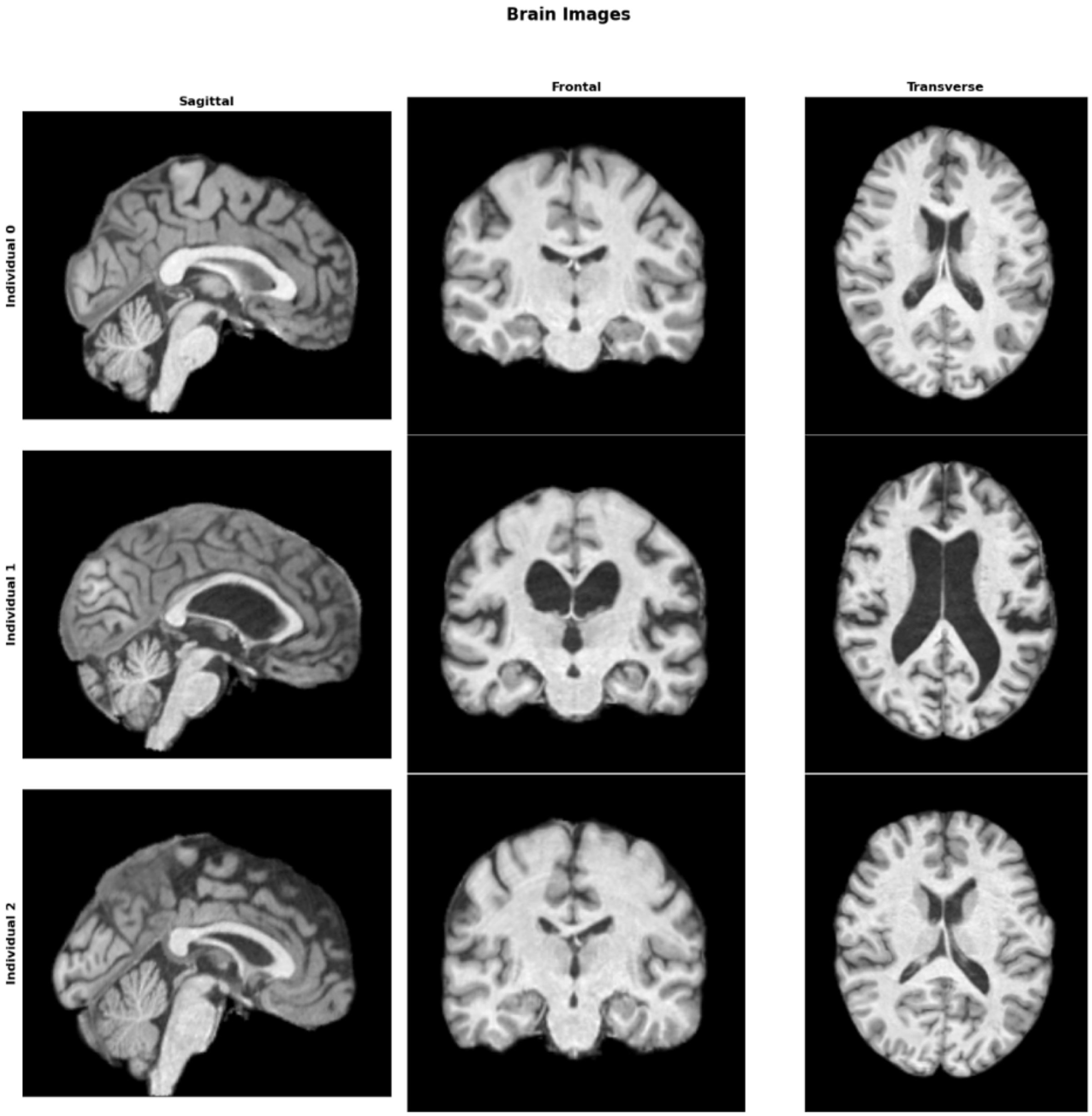


**Supplementary Figure 7 | Extreme outliers caused higher than average reconstruction loss.** Above brain scans show severe anomalies from three individuals potentially due to scan quality, including motion-induced scan disturbance (Individuals 0 and 2), abnormality in brain structure (Individual 1). Notably, sagittal view of scan from Individual 2 shows missing parts in lower section of the image.

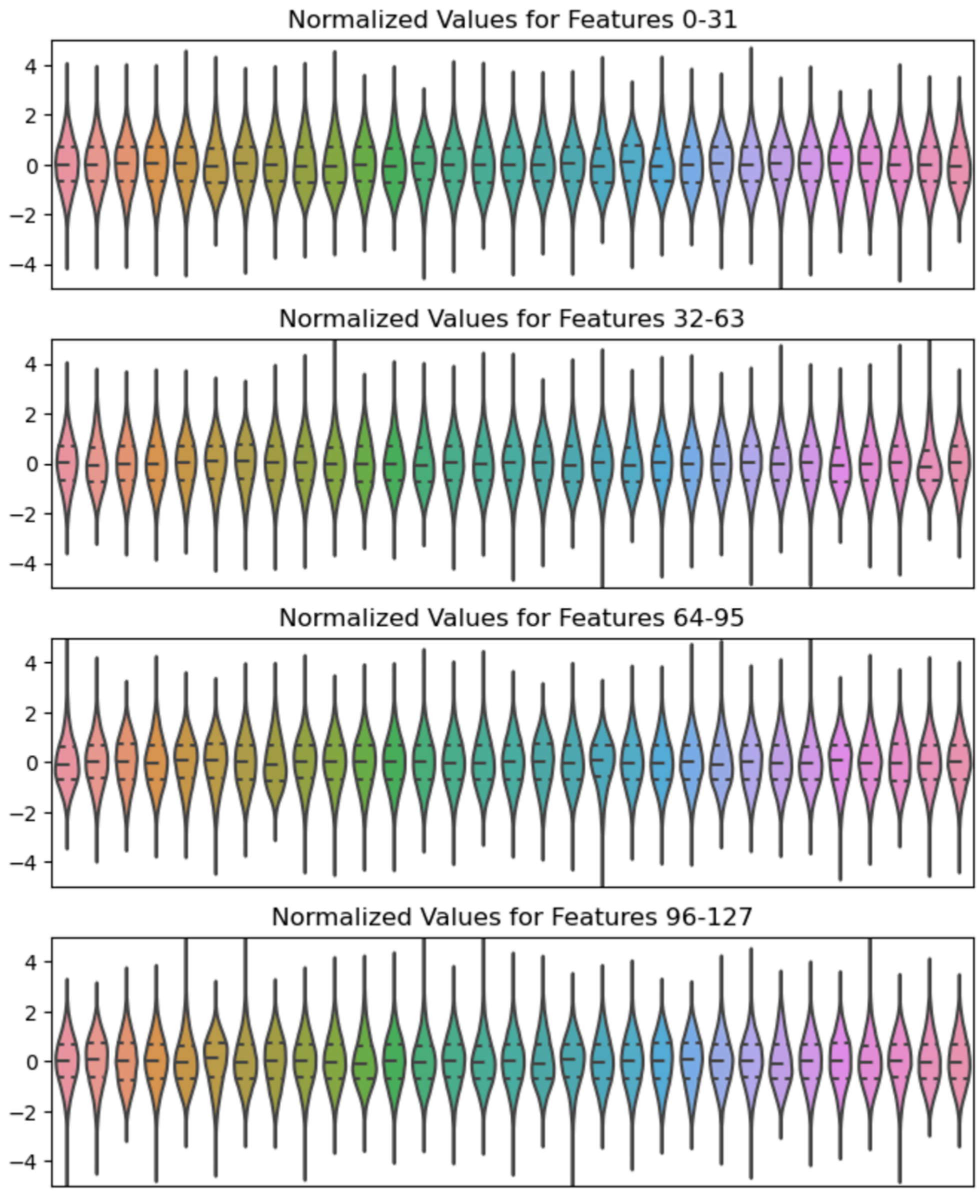


**Supplementary Figure 8 | Visualization of UDIPs showing normal distribution for each of the 128 dimensions upon inferencing.** Violin plots for UDIPs are shown for normalized distribution of each UDIP for entire testing dataset of 37,376 scans. Each plot has three lines: top line for upper quartile (Q3), middle line for median, lower line for lower quartile (Q1). Violin width represents kernel density estimate (KDE) of the data. The distributions demonstrate overall normality of the UDIPs.

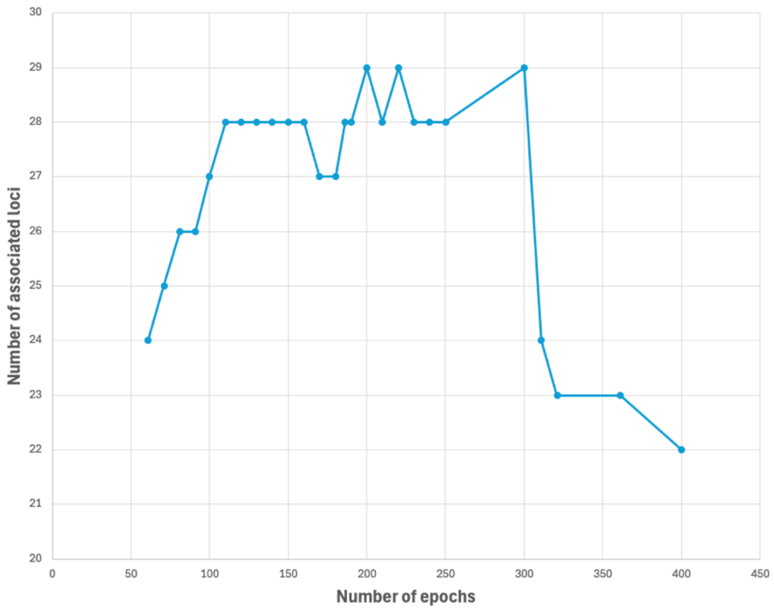


**Supplementary Figure 9 | Number of associated loci vs number of training epochs.** Model trained for 200, 220 and 300 epochs revealed the highest number of associated loci. Out of 29 loci, 10 were not found in previous CNN-based UDIP pipeline.

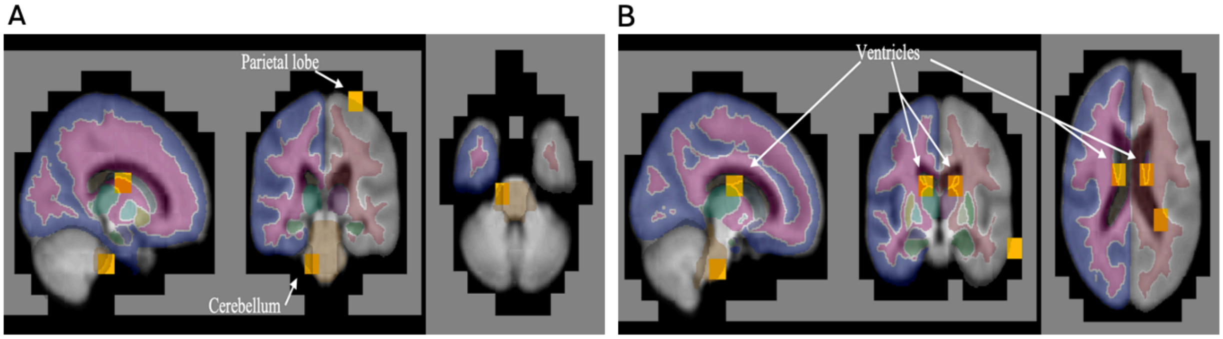


**Supplementary Figure 4 | Attention head analysis. A.** Attention heads revealed long-range regions from across the brain like structural regions of cerebellum and parietal lobe. **B.** Symmetrical regions were captured by ViT attention heads, such as left and right ventricles at the subcortical level.

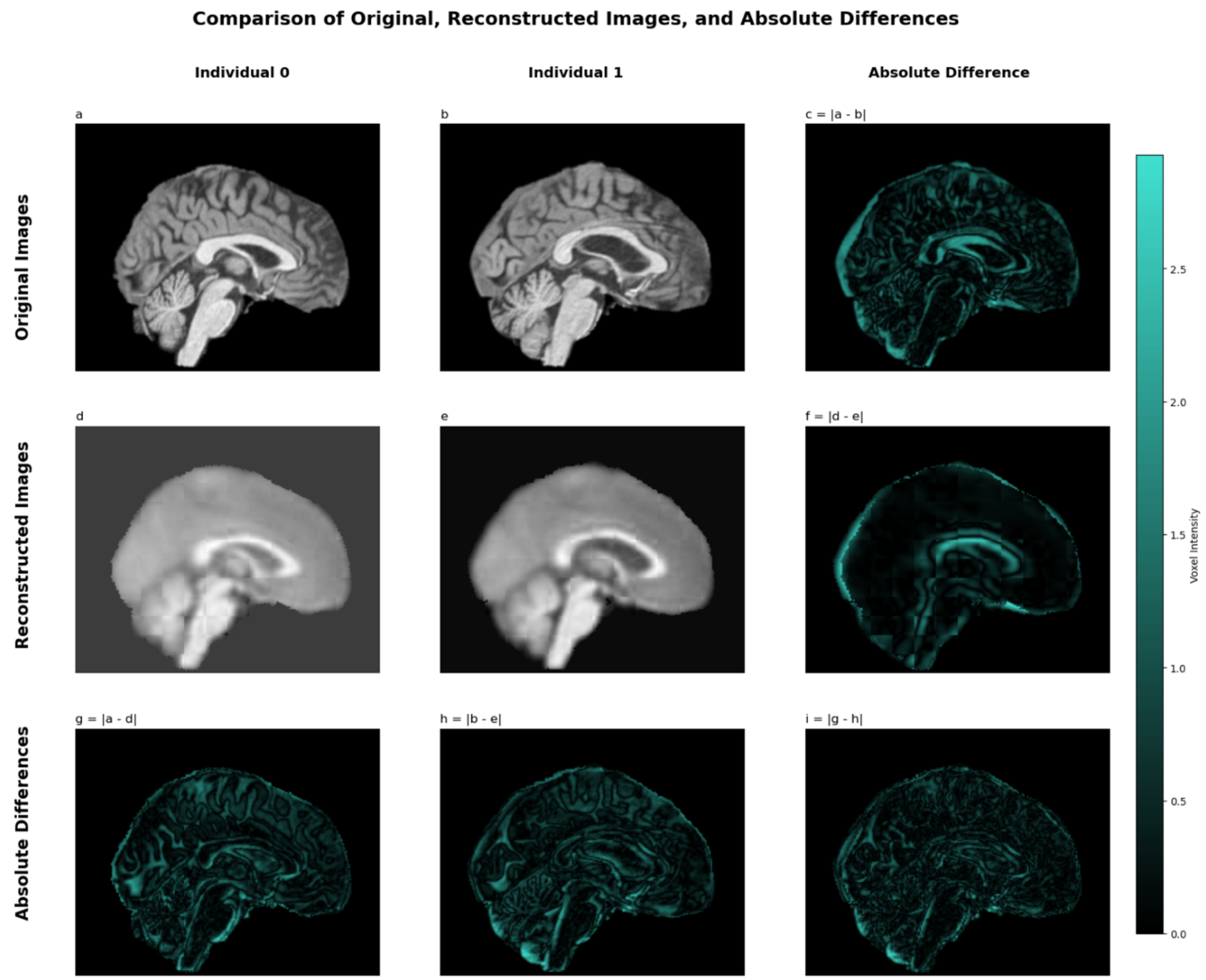


**Supplementary Figure 51 | Original, reconstructed and absolute differences between two individuals.** This figure shows reconstruction difference between Individual 0 and Individual 1. Additionally, differences between reconstructed and original images for Individual 0 and Individual 1 are compared.

### Supplementary Tables

| Loci ID | Lead SNP | Gene(s) |
|---|---|---|
| 1 | rs6658111 | RPL21P24, ATP6V0E1P4 |
| 2 | rs1609829 | KIAA1614 |
| 3 | rs4417739 | N/A |
| 4 | rs17279437 | SLC6A20 |
| 5 | rs4308324 | OSTN,GMNC |
| 6 | rs13107325 | SLC39A8 |
| 7 | rs4869744 | CCDC170 |

| | | |
|---|---|---|
| 8 | rs798546 | N/A |
| 9 | rs10272907 | N/A |
| 10 | rs3801382 | FAM3C |
| 11 | rs6973339 | LOC124901750 |
| 12 | rs4128399 | PRKAG2 |
| 13 | rs13250387 | N/A |
| 14 | rs2514524 | GDF6 |
| 15 | rs6469788 | TNFRSF11B |
| 16 | rs1409682 | LPAR1 |
| 17 | rs4978988 | N/A |
| 18 | rs35565319 | PAPPA |
| 19 | rs12253527 | MLLT10 |
| 20 | rs10770131 | IRAG1 |
| 21 | rs4074516 | LGR4 |
| 22 | rs7123402 | LINC02550 |
| 23 | rs74651308 | SLC01A2 |
| 24 | rs61920200 | CCDC91,PTHLH |
| 25 | rs12146713 | NUAK1 |
| 26 | rs2738265 | BMP4 |
| 27 | rs1441819 | N/A |
| 28 | rs4843553 | C16orf95 |
| 29 | rs224329 | GDF5-AS1,UQCC1 |

**Supplementary Table 1 | Loci discovered by ViT-AE GWAS**
